# Effect of the Choosing Wisely Canada campaign on prescription of nonsteroidal anti-inflammatory drugs in people with hypertension, heart failure and chronic kidney disease in Canada

**DOI:** 10.1101/2024.10.25.24316159

**Authors:** Celestin Hategeka, Lucy Cheng, Heather C Worthington, Tracey-Lea Laba, Michael R Law

## Abstract

The Choosing Wisely Canada (CWC) campaign focuses on helping physicians and patients engage in conversations about unnecessary tests and treatments. In 2014, a CWC recommendation advised against prescribing nonsteroidal anti-inflammatory drugs (NSAIDs) in individuals with hypertension, heart failure or chronic kidney disease (CKD). We evaluated the impact of this recommendation on prescription of NSAIDs in British Columbia (BC), Canada. We identified all persons continuously registered with BC’s Medical Service Plan at any point between October 1, 2011 and December 31, 2017 with hypertension, heart failure or CKD. Using prescription claims data and an interrupted time series analysis, we estimated the quarterly number of people initiating NSAIDs and the proportion of days covered (PDC). We also conducted sub-group analyses to study the impact of the recommendation by patients’ demographics (sex and age). We analyzed 1,479,704 NSAID claims from 903,732 patients with a diagnosis of hypertension, heart failure and/or CKD. Overall, we found no statistically significant change either immediately or over time in initiation and PDC of NSAIDs. However, we did observe a decrease in both initiation and PDC of NSAIDs over time among male patients and patients aged ≤65 years. We found that the CWC recommendation had a mixed impact, with initiation and PDC of NSAIDs significantly declining only among male and younger patients.

**Highlights:**

- Reducing low-value care can help to counter increasing healthcare spending.
- CWC can contribute to reducing low-value care, however, evidence on its effectiveness remains elusive.
- CWC led to NSAIDs prescription decline for patients ≤65 years and male patients.
- Overall, NSAID prescribing has been declining over time irrespective of CWC.

## INTRODUCTION

Globally, health spending has risen over the past few decades and currently represents approximately 10% of gross domestic product (GDP).^1^ Canada and the United States (US) spent approximately 10.4% and 17.2% of GPD, respectively, on health in 2017.^2^ It is expected that health spending will continue to increase given aging populations in these countries.^2^ At the same time, spending on low-value care—medical treatments or tests whose risk of harm exceed likely benefit or are not cost effective^3^—is a significant challenge and accounts for a considerable share of health spending. For example, as much as 30% of US health spending is attributed to low-value care.^4^ Similarly, a 2017 Canadian Institute for Health Information report revealed that up to 30% of patients were provided low-value care in Canada.^5^

Reducing low-value care has been advocated as one of the approaches to counter increasing health care spending while also improving the quality of health care.^5-8^ One effort to achieve this goal has been the Choosing Wisely (CW) campaign, which focuses on reducing or preventing *therapeutic illusion* by helping physicians and patients have conversations about unnecessary and potentially harmful tests, treatments, and procedures.^5-7, 9^ The campaign was first introduced in the US in 2012, and several other countries have followed, including Canada in 2014.^10, 11^ Currently, the CW Canada (CWC) campaign has over 330 recommendations developed by professional societies that represent different clinical specialties.^12^ Each medical professional society working with its members has identified a list of CWC recommendations concerning low-value care that physicians should stop /avoid and patients should question.^12^ Accordingly, the Canadian Society of Nephrology (CSN) developed its list of five CWC recommendations including “don’t prescribe nonsteroidal anti-inflammatory drugs (NSAIDs) in individuals with hypertension or heart failure or CKD of all causes, including diabetes”.^13-15^ The rationale behind this recommendation is that using NSAIDs, including cyclo-oxygenase type 2 inhibitors, can increase “blood pressure, make antihypertensive drugs less effective, cause fluid retention and worsen kidney function”.^14, 15^ This recommendation also highlights that other medications such as “acetaminophen, tramadol, or narcotic analgesics (short-term use), may be safer than and as effective as NSAIDs”.^14, 15^

Early findings on the impact of the CW campaign have been mixed.^4, 8^ For example, Rosenberg and colleagues assessed the impact of seven CW recommendations in US and found that only two (use of imaging for headache and cardiac imaging) declined following the campaign, while at the same time, the use of NSAIDs increased.^10^ In Canada, a 2018 study by Welk and colleagues evaluated the impact of the campaign targeting three Canadian Urological Association’s CWC recommendations, and found no significant change in physicians’ practice patterns in Ontario after the campaign.^16^ However, strong evidence on the impact of several other CWC recommendations, including not prescribing NSAIDs in selected conditions, in Canada is yet to be demonstrated. At the same time, recent estimates suggest that the proportion of patients that receive low-value care in Canada remains high (~30%), with significant variations across regions.^17^ In this study, we aimed to evaluate the impact of the CWC campaign on the prescription of NSAIDs among patients with hypertension, heart failure and CKD in British Columbia (BC), Canada.

## MATERIALS AND METHODS

### Design

We conducted a quasi-experimental interrupted time series (ITS) study to examine longitudinal changes in prescription of NSAIDs following the release of the CWC campaign while rigorously controlling for pre-existing trends ^29, 30^

### Setting and policy intervention

British Columbia is Canada’s third most populous province, with approximately 5 million people (median age of ~43 years).^18, 19^ The province has a single-payer Medical Services Plan (MSP) that covers all medically necessary services for residents. However, outpatient prescriptions are covered by a mix of private, public, and out-of-pocket payments. Every prescription dispensed in community pharmacies in BC is recorded in a provincial drug database (BC PharmaNet),^20^ making it possible to track physician prescribing behaviours using dispensed prescriptions and ultimately evaluate the potential impact of policies such as the CWC recommendations that target these behaviors. The CWC recommendation of “don’t prescribe NSAIDs in individuals with hypertension or heart failure or CKD of all causes, including diabetes” was released publicly in Canada on October 29, 2014. We are not aware of any active dissemination of this recommendation in BC beyond the public release. This recommendation was endorsed primarily by nephrologists and is one of five CWC recommendations developed by CSN.^15^ Further details about the process that CSN used to develop its five CWC recommendations has been discussed elsewhere.^15^

### Data sources

We used de-identified, linked data from the BC Ministry of Health and provided through Population Data BC, ^21^ covering the period between October 01 2011 and December 31 2017. Data linkage used patient name, unique patient health number, date of birth, and unique practitioner identifier. Overall, we used data that came from six databases. (1) The PharmaNet database provided data on all NSAID prescriptions dispensed in BC, including the Anatomical Therapeutic Classification code (ATC) and Drug Identification Numbers.^22^ PharmaNet does not capture information about medications dispensed individuals who are offered drug benefits through the Federal government (e.g. veterans, First Nations). (2) The MSP payment file provided data on outpatient services including patients’ diagnosis (hypertension or heart failure or CKD, in this study).^23^ (3) The Discharge Abstract Database (DAD) provided data on hospital discharge diagnoses including hypertension or heart failure or CKD.^24^ (4) The central registry file and (5) Vital Statistics Mortality database provided data on the population including age, sex, and date of death.^25,26^ (6) The College of Physicians and Surgeons of BC provided data on prescriber’s speciality.^27^

### Population

We identified all individuals diagnosed with hypertension, heart failure and / or CKD, and continuously registered with BC’s MSP between October 1, 2011 and December 31, 2017. Individuals who ceased enrolment in MSP for reasons other than death were excluded. Drawing on previous research,^28^ we narrowed our final study cohort to individuals who met at least one of the following criteria:

i) From the MSP payment information file, we identified all individuals with the following International Classification of Diseases (ICD)-9 diagnosis codes within the period indicated below between October 2011 and December 2017:

- ICD-9: 401-405, 428 (any two consecutive claims within a two-year period)
- ICD-9: 585 (only individuals with three or more claims within one-year period)

ii) From the DAD, we identified all individuals with one or more of the following ICD-10 diagnosis codes (ICD-10: I10-I15, I50, N18), in any diagnosis position, between October 2011 and December 2017.

### Outcomes

We used ATC code (M01) to identify prescriptions that include NSAIDs. We then studied the impact of the CWC recommendation on the following outcomes:

- *Proportion of days covered (PDC) for NSAIDs*: We calculated the ratio of the number of days in a quarter (three-month period) when NSAIDs were prescribed divided by the number of days in a quarter (i.e., quarterly PDC) per 1000 individuals continuously registered with MSP or until time of death. The lower the PDC for NSAIDs the less the exposure (or prescription) to NSAIDs.
- *Initiating NSAIDs:* We calculated the proportion of individuals initiating NSAIDs per quarter per 1000 individuals continuously registered with MSP or until time of death.

### Statistical Analysis

To fit our statistical models, we first calculated quarterly PDC for NSAIDs per 1000 individuals and quarterly proportion of individuals initiating NSAIDs per 1000 individuals across our datasets. We also weighted our estimates by the size of population to adjust for deaths in our study cohort. We then used these aggregated figures to fit segmented linear regression models, which had the following basic structures, with terms for (1) baseline intercept, (2) trend before the CWC recommendation release, (3) level change at time of the CWC recommendation release, and (4) trend change after the CWC recommendation release. Our parameters of interest were the coefficients for terms (3) and (4), representing any immediate changes in the level and quarterly trend change in our outcomes post-CWC recommendation, respectively, while controlling for pre-existing level and trend.

As quarterly observations may have been correlated over time, we assessed and controlled for autocorrelation as appropriate.^29^ We also allowed a phase in of one quarter because the CWC recommendation was released in the middle of the quarter, but also to allow enough time for the recommendation to potentially start being incorporated into the practice of prescribers. Additionally, we conducted subgroup analyses, determined a priori, to investigate whether the CWC recommendation had differential impact. Specifically, we stratified our analyses by patients’ demographics (sex and age). All analyses were conducted using SAS v 9.4 and R v 4.0.2. The study was approved by the University of British Columbia Behavioural Research Ethics Board (Certificate # H16-02087).

## RESULTS

### Cohort characteristics

Supplementary Appendix Table 1 describes the characteristics of our study cohort. We analyzed 1,479,704 NSAID claims from 903,732 patients with a diagnosis of hypertension (HTN), heart failure (HF) and/ or CKD, comprising 50.6% females and 56.1% aged over 65 years. Overall, 90.2% of the claims were prescribed by family doctors, while the prescriptions ordered by nephrologists (who were the primary endorser of the CWC recommendation) accounted for just 0.03%. Cardiologists, rheumatologists and other specialties accounted for 0.04%, 1.6% and 8.3%, respectively.

### Overall impact of the CWC recommendation

As shown in Figures 1 and 2, the trends in initiation (trend: −0.67/1000 / quarter, 95% CI: −0.87 to −0.43, p<0.001) and PDC of NSAIDs (trend: −0.39/1000/quarter, 95% CI: −0.45 to −0.34, p<0.001) were already decreasing before the CWC recommendation release. Following the release, we found no statistically significant change in either level or trend in proportion of people initiating NSAIDs (level: −0.61/1000, 95% CI: −2.86 to 1.63, p=0.59; trend: −0.22/1000/quarter, 95% CI: −0.53 to 0.08, p=0.16), neither was its PDC (level: 0.15/1000, 95% CI: −0.45 to 0.75, p=0.62; trend: −0.06/1000/quarter, 95% CI: −0.13 to 0.02, p=0.13) (Figures 1 and 2).

**Figure 1.**
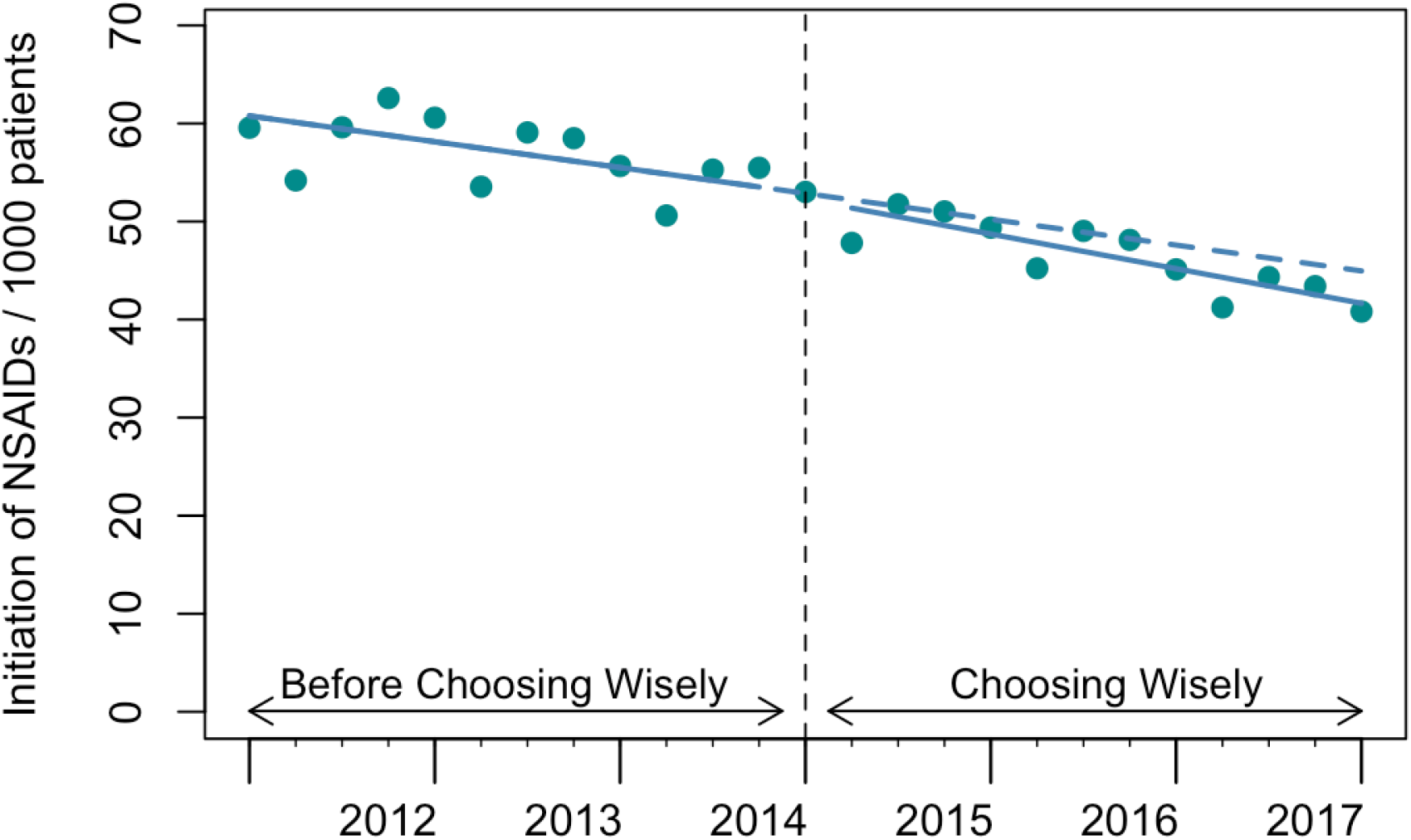
Quarterly time series of initiation of NSAIDs.

**Figure 2.**
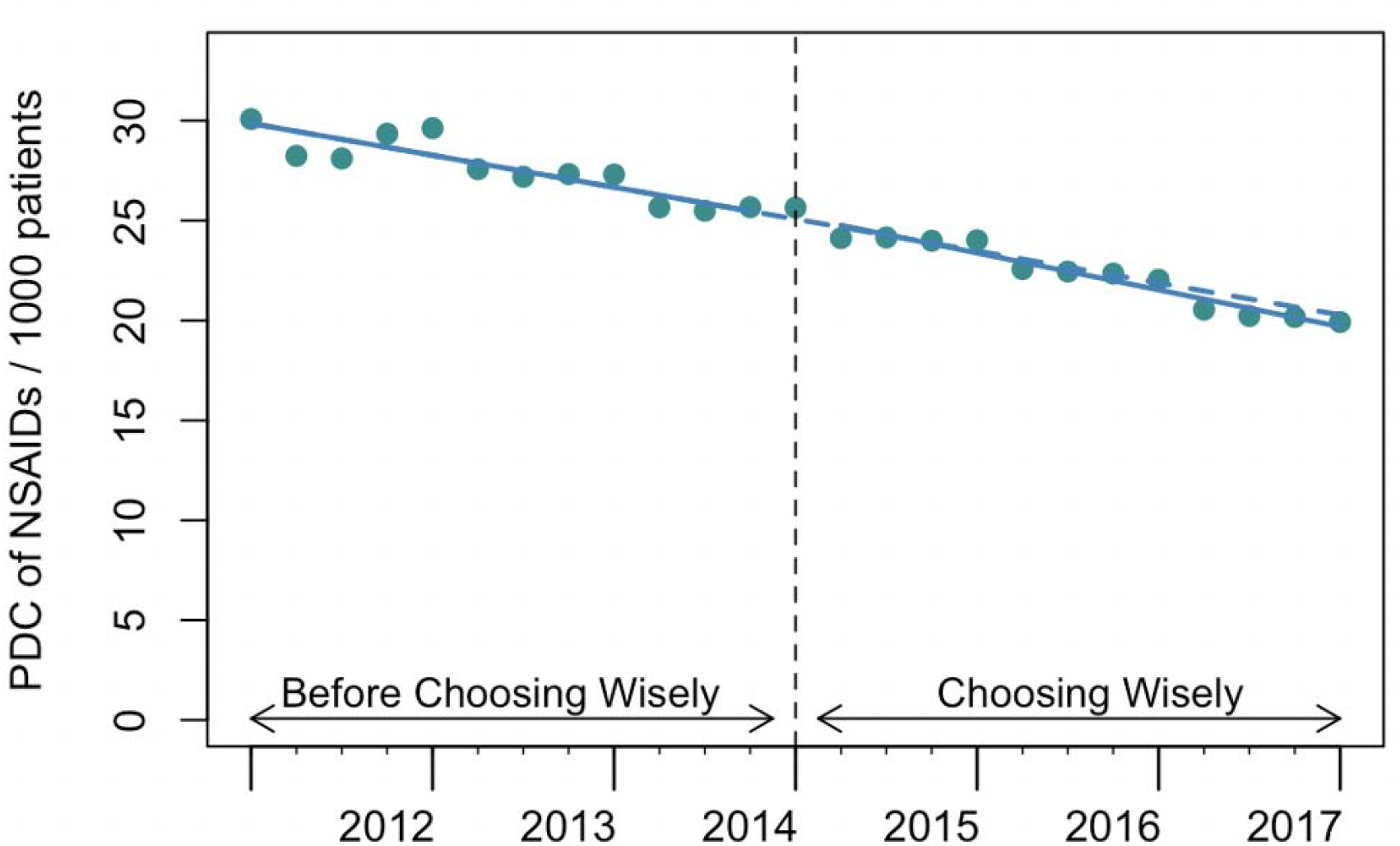
Quarterly time series of PDC of NSAIDs.

### Impact of the CWC recommendation by sex

The pre-existing trends in initiation and PDC of NSAIDs were decreasing significantly for female patients before the release of the CWC recommendation, as were the pre-existing trends for male patients (Figure 4 and Appendix Table 1). After the release of the CWC recommendation, we found no statistically significant change in either level or trend in initiation (level: −0.03 / 1000, 95% CI: −1.05 to 0.99, p=0.95; trend: −0.02 / 1000 / quarter, 95% CI: −0.16 to 0.13, p=0.82) and PDC (level: 0.16/1000, 95% CI: −0.08 to 0.42, p=0.21; trend: 0.01 /1000/ quarter, 95% CI: −0.02 to 0.04, p=0.54) of NSAIDs for female patients (Figure 3. A & C). However, we observed a statistically significant decline in the trend of initiation (trend: −0.20 / 1000 / quarter, 95% CI: −0.39 to −0.02, p=0.04) and PDC (trend: −0.07 / 1000 / quarter, 95% CI: −0.12 to −0.02, p=0.01) of NSAIDs for male patients (Figure 3. B & D). Cumulatively after three years, initiation and PDC of NSAIDs among male patients decreased by 13.5% and 10.1%, respectively, that would have been expected absence the CWC recommendation.

**Figure 3.**
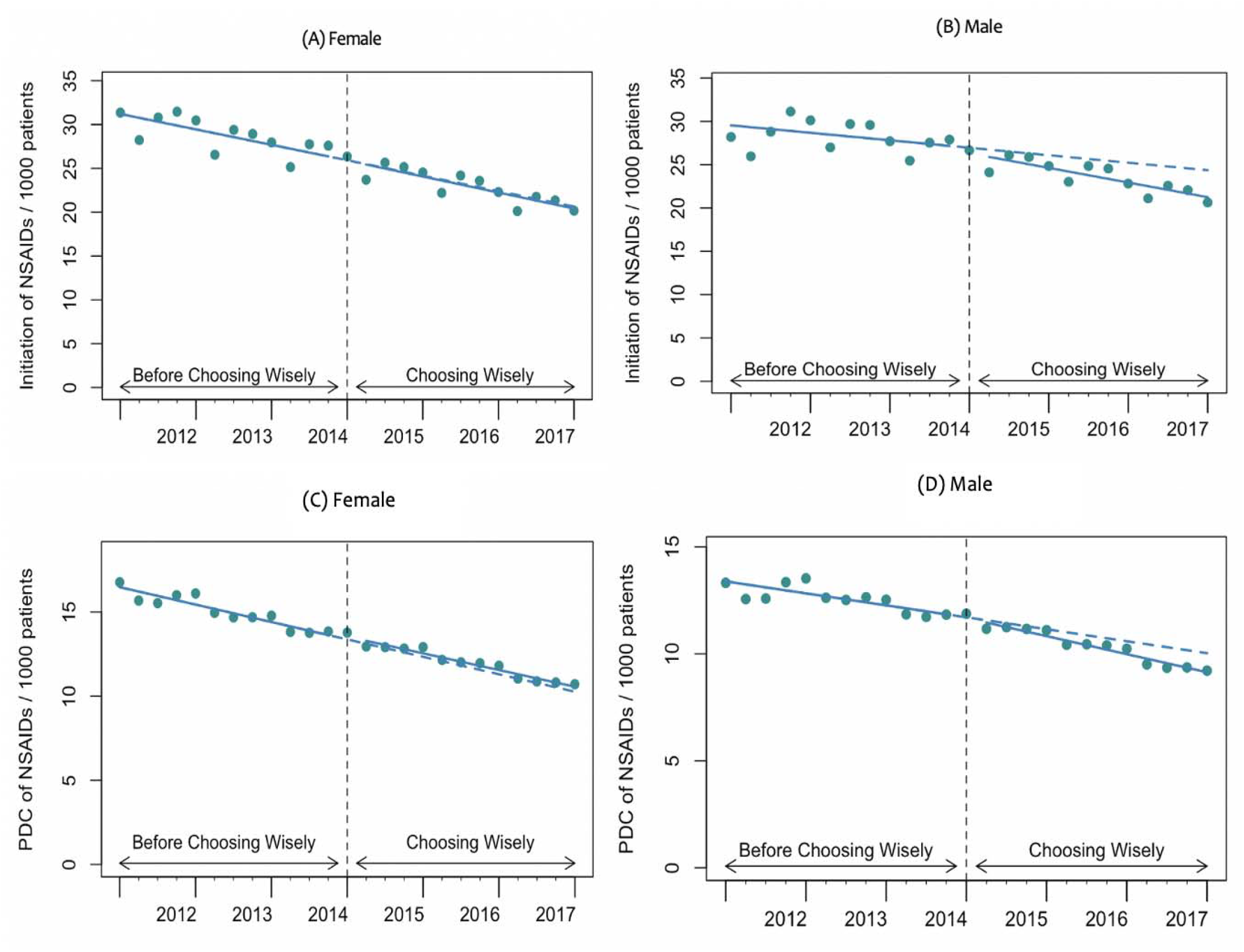
Quarterly time series of initiation and PDC of NSAIDs stratified by sex.

**Figure 4.**
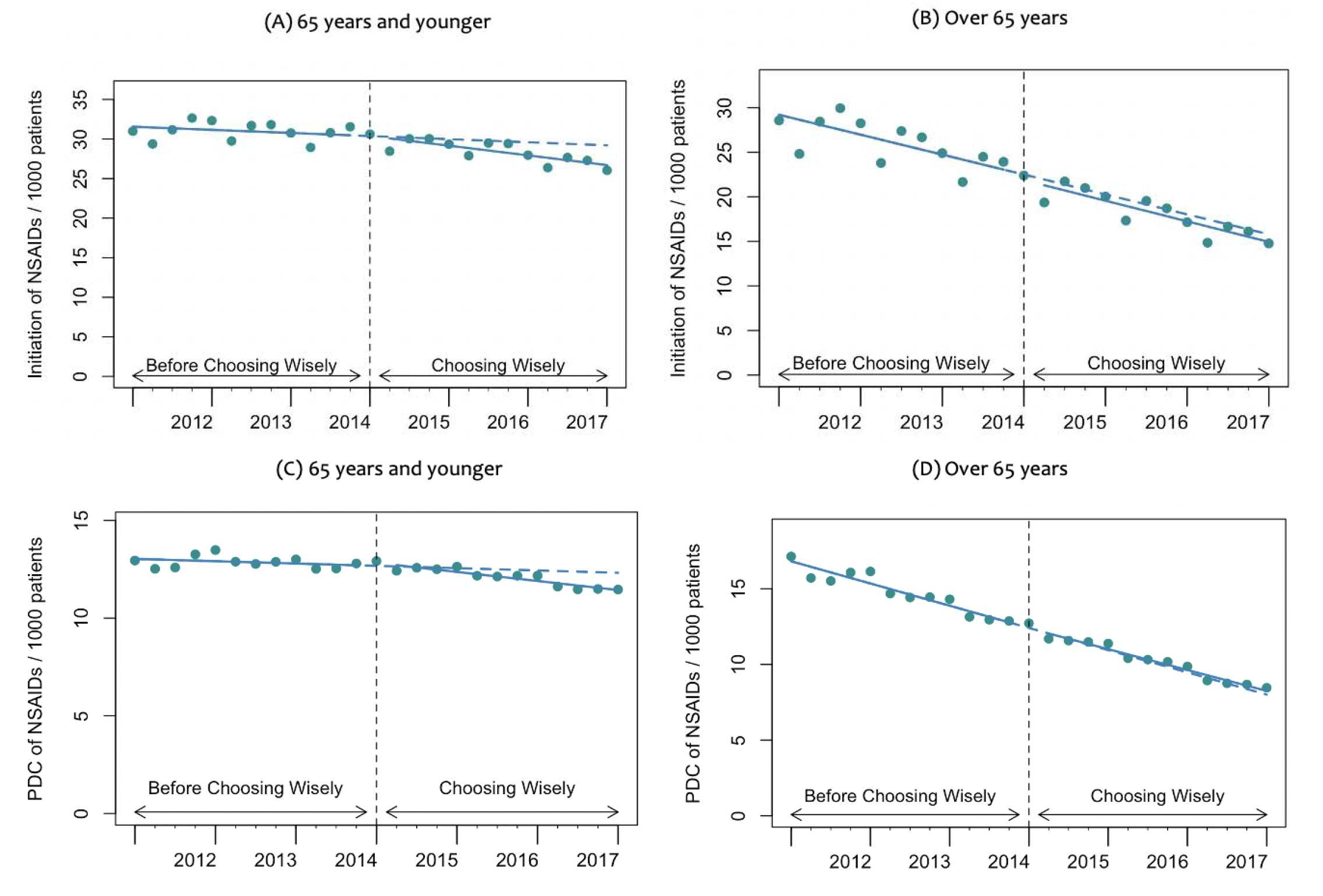
Quarterly time series of initiation and PDC stratified by age.

### Impact of the CWC recommendation by age

Our analyses stratified by age category (≤ 65 years vs > 65 years) showed that the pre-existing trends in initiation (trend: −0.56/1000/quarter, 95% CI: −0.69 to −0.42, p<0.001) and PDC (trend: −0.36/1000/quarter, 95% CI: −0.39 to −0.33, p<0.001) of NSAIDs were decreasing significantly for individuals older than 65 years in our cohort before the release of the CWC recommendation (Figure 4. B & D and Appendix Table 1). Similarly, the pre-existing trends in initiation (trend: −0.09/1000/quarter, 95% CI: −0.18 to −0.007, p=0.04) and PDC (trend: −0.03/1000/quarter, 95% CI: −0.06 to 0.001, p=0.07) of NSAIDs were also decreasing, but marginally, for the younger cohort (Figure 4. A & C). After the release of the CWC recommendation, we observed a modest, but statistically significant, trend decrease in both initiation (trend: −0.20/1000/quarter, 95% CI: −0.33 to −0.08, p=0.003) and PDC (trend: −0.08/1000/quarter, 95% CI: −0.12 to −0.04, p<0.001) of NSAIDs for the younger cohort (Figure 4. A & C). This represents, after three years, a reduction of 8.8% for initiation and 7.4% for PDC of NSAIDs in the younger cohort that would have been expected absence the recommendation. However, there was no statistically significant change in level and trend for both initiation and PDC of NSAIDs for the older cohort (Figure 4. B & D).

## DISCUSSION

We found that the CWC recommendation had a mixed impact on the use of NSAIDs among patients with hypertension, heart failure or CKD. In the overall cohort, the recommendation did not significantly change either the initiation or PDC of NSAIDs. However, subgroup analyses revealed some differential impact based on patients’ sex and age. In particularly, we found that the recommendation was associated with a significant decline in trend in both initiation and PDC of NSAIDs for patients ≤ 65 years and male patients. Furthermore, our analyses showed that while the recommendation was endorsed by nephrologists, most of the NSAID prescriptions were ordered by family doctors.

Our overall results appear to be consistent with previous studies evaluating the impact of the CWC campaign, suggesting the campaign has not had significant impact on reducing low-value care in Canada. For example, Welk and colleagues evaluated the impact of the campaign targeting three Canadian Urological Association’s CWC recommendations and found no significant change in prescribing behaviors in Ontario, Canada, after the campaign.^16^ Beyond Canada, however, our findings appear to contradict the Rosenberg and colleagues’ study on early impact of the CW campaign in US, that showed a significant increase in prescribing of NSAIDs in patients with hypertension, heart failure and / or CKD following the campaign.^10^ In our overall analyses, we found that the CWC campaign was associated with a modest significant decrease in trend in PDC and a non-statistically significant decrease in trend in initiation of NSAIDs in BC, Canada. Similarly, it should be noted that irrespective of the CWC campaign, the changing prescribing behaviors in BC appear overall to be in the right direction, i.e., decline in NSAIDs prescribing over time. The results of our sub-group analyses suggest that the CWC campaign had some differential impact, varying by patients’ sex and age. The differential impact appears to correlate with pre-existing trends before the CWC recommendation, with the campaign overall having a significant intended impact where the prescribing behaviors of NSAIDs had not significantly been leaning in the right direction. Specifically, the CWC campaign had a positive impact on trends in NSAID prescribing for both male and patients ≤ 65 years as their pre-existing trends had not been decreasing as fast as the observed decrease in pre-existing trends among female and patients older than 65 years (Figures 3 and 4).

There are several possible explanations for the overall limited impact of the CW campaign, including selection of the recommendations and implementation strategies.^8, 31, 32^ Some have argued that many CW recommendations are based on weak evidence, making their uptake challenging. However, the CWC recommendation we studied has a strong evidence-base, suggesting its successful uptake would improve outcomes for patients with the three targeted conditions,^15^ but our analyses show little evidence that it impacted care. Consistent with other interventions that just provide information to clinicians, CWC is unlikely to change physician behaviour meaningfully through a passive campaign. Evidence suggests that active interventions are often needed to change physician behaviour.^33^ Moreover, while it has been suggested that physicians are generally aware of the CWC recommendations, with over 330 CWC recommendations, physician saturation is likely and it is also difficult to tell if many were aware of the specific recommendation that we studied.^12, 34^ Similarly, less than a quarter (~22%) of the CSN members participated in the selection of the five CWC CSN recommendations, including the one we studied.^15^ A recent reflection on five years of the CWC campaign suggests that tackling physician, patient and health system level factors would be necessary for the campaign to significantly impact care in Canada.^8^

Our study has limitations that need to be acknowledged. First, given that the implementation of the CWC campaign may not have been homogenous across Canada, it is not clear if the campaign was as active in BC as other provinces or territories, so our results may not fully generalize across Canada. Second, our analytical cohort may not have been comprehensive given we relied solely on ICD codes to select the cohort members. Prior research suggests that identifying CKD using ICD codes in administrative data has low sensitivity (but very high specificity).^35^ Although using laboratory test and/or pathology results would enhance the sensitivity with respect to identification of CKD, these data are not available in the administrative data that we used. However, given this low sensitivity was likely consistent across our study period, we do not expect our results to be impacted.

In conclusion, we found that the CWC campaign had a mixed impact, with prescription of NSAIDs declining only among male and younger patients with hypertension, heart failure and / or CKD. While the recommendation we studied was endorsed by nephrologists, most of the NSAID prescriptions were generally ordered by family doctors, suggesting a room for future improvement should the recommendation focus on major prescribers of NSAIDs. Similarly, irrespective of the CWC campaign, the changing prescribing behaviors in BC, Canada, appear overall to be in the right direction, with NSAID prescribing declining over time.

## Data Availability

All data produced in the present work are contained in the manuscript

## ACKNOWLEDGEMENTS

This analysis was funded by a Foundation Scheme Grant from the Canadian Institutes for Health Research (FDN-148412, PI: Dr. Law). Dr. Hategeka received support through a Vanier Canada Graduate Scholarship and a Banting Postdoctoral Fellowship from the Canadian Institutes of Health Research. Dr. Law received salary support through a Canada Research Chair in Access to Medicines and a Michael Smith Foundation for Health Research Scholar Award. Findings presented during the 72nd Annual Scientific Session of the American College of Cardiology Together With WCC (ACC.23/WCC), March 4-6, 2023, live in New Orleans, LA.

## DISCLAIMER

Data for this study were obtained via Population Data BC, including data from the BC PharmaNet system. All inferences, opinions and conclusions drawn in this publication are those of the authors and do not reflect the opinions of Population Data BC or the data steward(s).

