## Supplemental file CW for "Effect of the Choosing Wisely Canada campaign on prescription of nonsteroidal anti-inflammatory drugs in people with hypertension, heart failure and chronic kidney disease in Canada": Supplementary Appendix .docx

Table 1: Characteristics of the study cohort

| Variable | Cohort | Percent | HTN | Percent | HF | Percent | CKD | Percent |
| --- | --- | --- | --- | --- | --- | --- | --- | --- |
| N^†^ | 903,732 | 100.00 | 856,704 | 94.8 | 143,104 | 15.8 | 91,621 | 10.1 |
| Female | 457,169 | 50.6 | 434,785 | 50.8 | 68,561 | 47.9 | 45,534 | 49.7 |
| Male | 446,563 | 49.4 | 421,919 | 49.3 | 74,543 | 52.1 | 46,087 | 50.3 |
| <=65 years | 395,656 | 43.8 | 381,852 | 44.6 | 23,154 | 16.2 | 19,055 | 20.8 |
| Over 65 years | 508,076 | 56.2 | 474,852 | 55.4 | 119,950 | 83.8 | 72,566 | 79.2 |

^†^ A combination of any of these three diseases is possible for a single patient (that is why adding % up leads to > 100%).

HTN, hypertension; HF, heart failure; CKD, chronic kidney disease.

Table S1. Results of segmented regression analysis of effect of CWC on NSAIDs prescription in BC, Canada

|  | Intercept  (95% CI) | Pre-existing trend  (95% CI), p-value | Level change  (95% CI), p-value | Trend change  (95% CI), p-value |
| --- | --- | --- | --- | --- |
| Overall cohort |  |  |  |  |
| - Initiation | 61.42/1000  (59.82 to 63.02) | -0.65/1000  (-0.87 to -0.43), p<0.001 | -0.61/1000  (-2.86 to 1.63), p=0.59 | -0.22/1000  (-0.53 to 0.08), p=0.16 |
| - PDC | 30.26/1000  (29.84 to 30.68) | -0.39 /1000  (-0.45 to -0.34), p<0.001 | 0.15 / 1000  (-0.45 to 0.75), p=0.62 | -0.06 / 1000  (-0.13 to 0.016), p=0.13 |
| Sub-group analysis |  |  |  |  |
| *Sex* |  |  |  |  |
| *Female* |  |  |  |  |
| - Initiation | 31.64 /1000  (30.90 to 32.38) | -0.44/1000  (-0.54 to -0.34), p<0.001 | -0.03 / 1000,  ( -1.05 to 0.99), p=0.95 | -0.02 / 1000  (-0.16 to 0.13), p=0.82 |
| - PDC | 16.73/1000  (16.55 to 16.91) | -0.25/1000  (-0.28 to -0.23), p<0.001 | 0.16/1000  (-0.08 to 0.42), p=0.21 | 0.01 /1000  (-0.02 to 0.044), p=0.54 |
| Male |  |  |  |  |
| - Initiation | 29.75/1000  (28.76 to 30.74) | -0.21/1000  (-0.35 to -0.08), p=0.005 | -0.63/1000  (-2.05 to 0.79), p=0.39 | -0.20 / 1000  (-0.39 to -0.02), p=0.04 |
| - PDC | 13.52/1000  (13.25 to 13.79) | -0.14/1000  (-0.17 to -0.10), p<0.001 | -0.03/1000  (-0.43 to 0.36), p=0.86 | -0.07 / 1000  (-0.12 to -0.02), p=0.01 |
| *Age* |  |  |  |  |
| *≤ 65 years* |  |  |  |  |
| - Initiation | 31.64/1000  (30.99 to 32.30) | -0.097/1000  (-0.18 to -0.007), p=0.04 | 0.009/1000  (-0.92 to 0.94), p=0.98 | -0.20/1000  (-0.33 to -0.08), p=0.003 |
| - PDC | 13.06 /1000  (12.83 to 13.28) | -0.029/1000  (-0.06 to 0.001), p=0.07 | 0.15/1000  (-0.17 to 0.48), p=0.37 | -0.08/1000  (-0.12 to -0.04), p=0.0005 |
| *>65 years* |  |  |  |  |
| - Initiation | 29.76/1000  (28.78 to 30.74) | -0.55 /1000  (-0.69 to -0.42), p<0.001 | -0.62/1000  (-2.00 to 0.74), p=0.38 | -0.018/1000  (-0.20 to 0.1704761), p=0.84 |
| - PDC | 17.18/1000  (16.96 to 17.40) | -0.36/1000  (-0.39 to -0.33), p<0.001 | -0.024/1000  (-0.33 to 0.28), p=0.87 | 0.022/1000  (-0.018 to 0.06), p=0.29 |

PDC, proportion of days covered; CI, confidence interval.
